# Modifying gut integrity and microbiome in children with severe acute malnutrition using legume-based feeds (MIMBLE II): A Phase II trial

**DOI:** 10.1101/2023.05.29.23290673

**Authors:** Kevin Walsh, Agklinta Kiosia, Peter Olupot-Oupot, William Okiror, Tonny Ssenyond, Charles Bernard Okalebo, Rita Muhindo, Ayub Mpoya, Elizabeth C George, Gary Frost, Kathryn Maitland

**Affiliations:** Division of Diabetes, Endocrinology and Metabolism, Imperial College, 6th Floor Commonwealth Building, Hammersmith Campus, DuCane Road, London W12, UK; Dept. of Nutritional Sciences. School of Life Course & Population Sciences, Faculty of Life Sciences & Medicine, King’s College, London SE1 9NH, UK; Mbale Clinical Research Institute, Busitema University Faculty of Health Sciences, Mbale Campus, Palissa Road, PO Box 1966, Mbale, Uganda; Kenya Medical Research Institute (KEMRI)-Wellcome Trust Research Programme, Kilifi, Kenya; Medical Research Council Clinical Trials Unit (MRC CTU) at University College London, UK; Imperial College, Department of Infectious Disease and Institute of Global Health and Innovation, Faculty of Medicine, Imperial College, London, UK

**Keywords:** severe malnutrition, mortality, randomised clinical trial, legume-base feeds

## Abstract

**Background:** Children hospitalised with severe malnutrition (SM) have high mortality and relapse/readmission rates. Current milk-based formulations targets restoring ponderal growth but not modification of gut barrier integrety or microbiome which increase risk of gram-negative sepsis and poor outcomes.

**Objectives:** We propose that legume-based feeds rich in fermentable carbohydrates will promote better gut health and improve overall outcomes.

**Methods:** We conducted at Mbale Regional Referral Hospital, Uganda an open-label Phase II trial involving 160 Ugandan children with SM (mid-upper arm circumference (MUAC) <11.5cm and/or nutritional oedema). Children were randomised to a lactose-free, chickpea-enriched legume paste feed (LF) (n=80) versus WHO standard F75/F100 feeds (n=80). Co-primary outcomes were change in MUAC and mortality to Day 90. Secondary outcomes included weight gain (>5 g/kg/day), *de novo* development of diarrhoea, time to diarrhoea and oedema resolution.

**Findings:** Increase in Day 90 MUAC was similar in LF and WHO arms (1.1 cm (interquatile range.IQR 1.1) vs 1.4cm (IQR 1.40) p=0.09. Day 90 mortality was similar 11/80 (13.8%) vs 12/80 (15%) respectively OR 0.91 (0.40 -2.07) p=0.83. There were no differences in any of the other secondary outcomes. Owing to initial poor palatability of the legume feed 10 children switched to WHO feeds. Per protocol analysis indicated a non-significant trend to lower Day 90 mortality and readmission rates in the legume feed (6/60: (10%) and (2/60: 3%) vs WHO feeds (12/71: 17.5%) and (4/71: 6%) respectively.

**Conclusion:** Further refinement of legume feeds and clinical trials are warrented given the poor outcomes in children with severe malnutrition.

**Trial registration:** ISRCTN 10309022.

**Funding:** **Confidence in Concepts – Joint Translational Fund 2017** (Imperial College, London)

## Introduction

Severe malnutrition (SM) remains a frequent cause of hospitalisation in African children. It is associated with high in-hospital mortality rates of ∼20%^1,2^ and poor long-term outcomes^3,4^. Even in the context of clinical trials addressing infection prophylaxis^3^ or modification of the WHO feed^5^ failed to improve the poor outcomes. Milk-based feeds recommended for management of SM (called F75 and F100) result in nutritional (anthropometric) recovery in survivors (the current gold standard of success) but this poorly predicts short and long term outcomes^6^, including increased risk of life-threatening events (death and/or re-hospitalisation with pneumonia or diarrhoea) in the 12 months following initial admission^4,7^. A Phase II trial examining other formulations compared a feed with reduced lactose and carbohydrate load in the starter feed compared to standard formula (F75). This did not demonstrate improvement in outcomes indicating more radical approaches are required in the design of nutritional feeds^5^.

There are multiple lines of evidence indicating that several domains of gut function are aberrant in children with SM. Intestinal atrophy^8,9^ results in functional loss of brush border disaccharidases (lactase, maltase and sucrase)^10,11^ which exacerbates diarrhoea and impairs recovery. Moreover, there is a significant relative microbiota immaturity and high levels of pathogenic flora in children with SM which are only partially ameliorated following three weeks of standard nutritional interventions^12^. We hypothesized that intestinal mucosal integrity and gut microbial diversity can be restored in SM by providing substrates that and induce fermentation in the gastrointestinal tract^13^. Fermentable carbohydrates can improve the balance of normal gut microbes and positively influence the immunological and metabolic function of the gut ^14,15^. Carbohydrates that escape digestion in the gastrointestinal tract (resistant starch and dietary fibre) induce favourable changes in colonic microbiota fermentation ^16^. These lead to the generation of short-chain fatty acids (SCFA) which have a positive influence on gut integrity and nutritional health by improving energy yield, modulation of colonic pH, production of vitamins and the stimulation of gut homeostasis, including anti-pathogen activities ^17,18^. We tested this hypothesis in a pilot trial (Modifying Intestinal Microbiome by Legume-Based fEeds: MIMBLE 1 PACTR201805003381361) which compared cowpea-supplemented standard nutritional formulae to standard WHO formuala (F75/F100)^19^. We demonstrated the feed was safe, palatable and resulted in equivalent weight and MUAC gain compared to standard WHO formulae (F75/F100)^19^. In the standard WHO feed arm faecal microbiota diversity showed very little change over the 28-day intervention nor change in major phyla. Furthermore, the SCFA concentrations on admission were approximately a third of the concentration of those reported in healthy African infants ^20^. However, in standard WHO feed arm, but not the cowpea arm there was a suppression of the SCFA propionate and butyrate at day 7 (to about 1/10^th^ of the normal concentrations) a period when the children are at high risk of mortality. We suspect that the suppression of SCFA at day 7 may have been due to the use of antibiotics, which recovered once antibiotic treatments were stopped. In-vitro batch culture (in an artificial colon) of the WHO milk feed (F75/100) demonstrated no impact on the gut microbiome or microbiologcal diversity whereas in the cowpea-enhanced feeds lead to increases in bifidobacteria (that has been linked to improved epithelial integrity^21^) and diversity. Since there were no differences on diarrhoea (frequency and resolution) or other clinical endpoints between the feeds, we made further modifications to the feed and developed, with a UK food manufacturer, a lactose-free, fermentable carbohydrate-containing alternative feed (Mimble 2 feed)^22^ swapping cowpea for chickpea. Here we report the Phase II Clinical Trial which compared a chickpea-supplemented lactose-free feed to standard milk feeds on a range of endpoints (MIMBLE 2). The trial was registered with **ISRCTN 10309022** on 23/05/2018.

## Methods

Modifying Intestinal Integrity and Microbiome in Malnutrition with Legume-Based Feed 2 [MIMBLE 2] was a single-centred (Mbale Regional Referral Hospital) open-label, proof-of-principle randomised controlled trial evaluating safety and efficacy of lactose-free chickpea based nutritional formulae compared to standard milk-based feeds.

### Screening, Randomisation and blinding

Children with suspected SM were clinically assessed for eligibility and exclusion criteria. Children aged 6 months to 12 years hospitalised with SM were eligible for inclusion in the trial enrolment. SM was defined as either marasmus (defined by mid-upper arm circumference (MUAC) <11.5cm) and/or kwashiorkor (defined as symmetrical pitting oedema involving at least the feet irrespective of weight for height Z score (WHZ) or MUAC) or a combination of both. Where prior written consent from parents/legal guardians could not be obtained, ethics committees approved parental verbal assent and deferred written informed consent as soon as practicable^23^. Otherwise, informed written consent was obtained from parents or guardians before randomisation. Children with SM with a comorbidity at very high risk of death e.g. malignant disease or terminal illness, or a parent/guardian not willing to consent were excluded from the trial.

An independent data manager, based at KEMRI-Wellcome Trust Research Programme (KWTRP), Kenya generated the sequential randomization list, using permuted blocks. This sequence was used, by a study administrator at KWTRP, Kilifi, Kenya, to prepare randomisation cards with the treatment allocation which were sequentially numbered and sealed in opaque envelopes, each signed across the seal ensuring allocation concealment. In the hospitals in Uganda randomisation was done by the study clinician using the numbered envelopes sequentially which contained the randomised feed strategy. Nurses/doctors were unblinded to study intervention; laboratory tests were assayed blinded. Children were randomly assigned 1:1 to either legume-based paste feed (investigational) or F75/F100 feeds (control) recommended by WHO.

### Study procedures

Children were managed on general pediatric ward. A structured clinical record documented relevant clinical, examination and laboratory baseline assessment. Nutritional feeds were given per protocol (the development of the feed^22^ recipe and feeding protocol is detailed in the published protocol)^24^. Briefly, for those in the control arm (WHO feeds) initially 130 ml/kg/day F75 therapeutic milk was given at 4-hourly intervals over the day until the child was stabilized and demonstrating appetite. At this point they transitioned to 4-hourly F100 therapeutic milk at the same rate and increased by 10 ml per feed, until a maximum rate of 200 ml/kg/day was achieved. Legume feeds were provided as a paste 4-hourly at 45-50g/kg/day (35-40g/kg/day if oedematous). With additional water per feed provided starting at 105-110ml/kg/day. Feed weight and additional water volume was adjusted daily in accordance to increasing weight (see supplemental material –feed volume/weight calculation chart). Once clinically stable the feed weight increased by 5g/feed until a maximum of 100g/kg/day. Mineral mix was added to the water for children in legume arm (as WHO milks already contain mineral mix). Thus, the quantity of legume feed provided matched the total amounts of energy and protein that would be received in the control arm, and additional water provided to match the fluid received. If the child took less than 80% of feed volume/weight for two consecutive feeds, despite attempts with spoon or syringe, then children were offered nasogastric tube feeding. Children in the legume strategy who could not initially tolerate non-fluid diet were switched to the WHO standard F75 feed and could then return to the legume strategy when non-liquid feeds were tolerated. All feed volumes and problems with feeding were recorded on standard proforma. Other standard treatments were prescribed including anti-malarials and antibiotics, following national guidelines.

Children were reviewed twice daily to discharge (generally ∼day 14). On consenting for the MIMBLE 2 study patients/parents/guardians agreed to remain in hospital for a minimum of 7 days but preferably 14 days (based on both the WHO and Ugandan Ministry of Health guidelines). Patients were permitted to leave earlier if they had no oedema if applicable, good weight gain and MUAC > 12.5cm. Serious adverse events (SAE) were actively solicited. Children were reviewed for clinical status and anthropometric status at 28- and 90-days post-randomisation. In addition, at admission, days 28 and 90.

### Endpoints

The co-primary endpoint was change in mid-upper arm circumference (MUAC) at Day 90 and mortality at Day 90. Secondary outcome measures include change of weight and achieving a weight gain of >5g/kg/day by day 28 and day 90, denovo development of diarrhoea (> 3 loose stools/day) and time to resolution of diarrhoea, time to oedema resolution and presence of oedema at days 28 and 90; and the number of serious adverse events (readmission to hospital to Day 90).

### Ethics

The ethics committees of Imperial College London, UK (**17IC4146)** and Mbale Hospital Research Ethics Committee, Uganda (**019/2018**) approved the protocol.

### Statistical Anaysis

Clinical data were analysed by using IBM Statistical Package for Social Sciences (SPSS) for Windows version 28 (Armonk, NY: IBM Corp) and R and R studio Core Team (2022). Calculation of weight for height (WHZ) was performed by using the online WHO Anthro Survey Analyser. Primary and secondary outcomes were analysed on an intention-to-treat and per-protocol basis. Per protocol analysis was defined as the analysis that included children that received and successfully completed their allocated treatment upon randomisation, during their hospital stay, and their survival status was known until study discharge (Day 90).

Statistical differences in baseline anthropometric and clinical characteristics between the two groups were assessed with the non-parametric Mann-Whitney U test and chi-square analysis for categorical data such as diarrhoea and oedema status from baseline and then on Days 7, 28 and 90. Cox regression analysis and Kaplan-Meier survival curves were employed to assess mortality and recovery differences between treatments, with competing risks regression analysis for readmissions, employing the Fine and Grey competing risk regression model, with the competing risk of death. Children were censored when abscoded or lost to follow up and in addition, children in the per protocol analysis were censored on the day that they had their allocated treatment switched and on survival analysis children were additionally censored when absconded or lost to follow-up.

Diarrhoea and oedema resolution from baseline to day 90 was also assessed via cox regression analysis. Results were identified as statistically significant when p<0.05 at 95% confidence interval (CI). Finally, to address missing data concerning lost to follow-up and absconded cases, multiple imputations (MI) analysis under the missing at random assumption, using predictive mean matching (PMM) was employed on MUAC, Weight, WGV, WHZ, oedema and diarrhoea status. The variables in the imputation model along with the number of values imputed are described in **Table S6** in supplemental file.

### Sample-size estimation

The overall sample size was 160 children (80 per each study arm). A formal sample size was not calculated as the aim was to generate adequate data of a proof of principal that the modified nutritional feed provides clinical, physiological and biological evidence of benefit to the child in terms of nutritional rehabilitation. Mid-upper arm circumference (MUAC) was selected as the primary criterion for nutritional recovery because it predicts mortality better and is less affected by oedema than other anthropometric measures^26^. Whilst a formal sample size was not calculated we were guided (for our primary endpoint) by a trial of antimicrobial prophylaxis, where in Kenyan children admitted with severe malnutrition the baseline mean MUAC was 10.6cm (SD 1.06) and by at 90 days 12.2cm (SD 1.35); a mean change of 1.6cm (SD 1.1) nutritional recovery at 90 days^3^.

## Results

Between 5^th^ July 2018 and 28^th^ August 2019 160 children, of a median age 17 (interquartile range, IQR 12-24) months were randomised to legume-based feed (n=80) or WHO feeds (n=80) and all children are included in the intention to treat analysis (**Figure 1**). Two (2.5%) and four (5%) of children in the legume and WHO arms respectively self-discharged (absconded) from hospital during the intial admission. In the legume and WHO arm 9 (11%) and 7 (9%) respectively were lost to follow up (survival status at 90 days unknown). One child (LF arm) withdrew from the trial. (**Figure 1** and **Table S1**)

**Figure 1.**
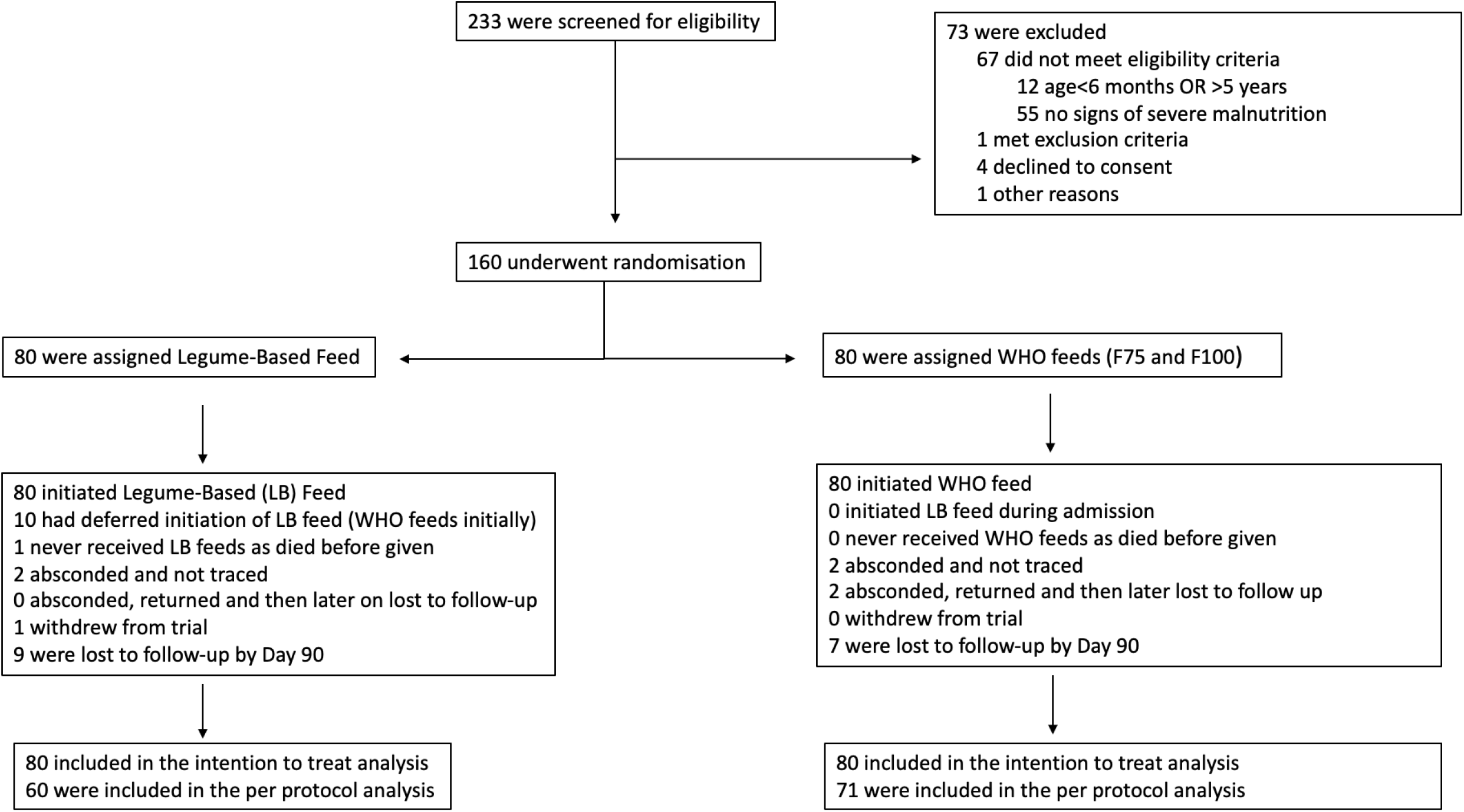
Trial Flow.

Baseline characteristics were balanced between randomized groups, except oedematous malnutrition was marginally more common in the legume feed (61% vs 53%) and of greater severity with 10/49 (20%) vs 6/41 (15%) respectively presenting with generalised oedema (**Table 1**). Overall, diarrhoea was present in 42 (26%) children but respiratory distress 6 (4%) and HIV were uncommon 7 (4%) as was pre-exisiting developmental delay 9(6%). Biochemical markers of severity (severe hyponatraemia and hypokalaemia) were present in 30 (19%) and 18 (11%) children respectively. Many had received antimicrobials prior to admission.

**Table 1:** Baseline Characteristics.

|  | **Legume Based**<br>**feed** | **WHO feeds**<br>**(F75/F100)** | **Total** |
| --- | --- | --- | --- |
| Participants, n | 80 | 80 | 160 |
| Age in months [Interquartile range] | 18 [12,24.7] | 17 [12,23.7] | 17 [12,24] |
| Sex: Male (%) | 44 (55) | 39 (48.75) | 83 (51.87) |
| **Nutritional status and history** |  |  |  |
| Mid-upper arm circumference (MUAC), cm [IQR] | 11.4 [10.5,12.2] | 11.2 [10.5,12.2] | 11.2 [10.5,12.2] |
| Weight for height Z score [IQR] | -3.79 [-4.4,-2.7] | -4.08 [-5,-2] | -4.0 [-4.6,-2.9] |
| Weight-for-height/length z score <-3 | 31 (39) | 39 (49) | 70 (44) |
| Oedema (kwashiorkor) | 49 (61) | 41 (51) | 90 (56) |
| Oedema severity: severe/generalized | 10/49 (20) | 6/41 (15) | 16/90 (18) |
| Signs of desquamation or flaky paint skin | 20/80 (25) | 15/80 (19) | 35/160 (22) |
| Age when feeds introduced (months) | 4 [3,6] | 5 [3,6] | 5 [3,6] |
| Currently breast feeding | 21/80 (26.5) | 24/80 (30) | 45/160 (28) |
| Previous admission with severe malnutrition | 5/80 (6) | 4/80 (5) | 9/160 (6) |
| **Complications at Presentation** |  |  |  |
| History of fever | 63/80 (79) | 57/80 (71) | 120/160 (75) |
| Fever (axillary temp) > 37.5°C | 10/80 (12.5) | 10/80 (12.5) | 20/160 (12.5) |
| Cough | 59/80 (74) | 59/80 (74) | 118/160 (46) |
| Indrawing or deep breathing | 3/80 (4) | 3/80 (4) | 6/160 (4) |
| Vomiting | 23/80 (29) | 25/80 (31) | 48/160 (19) |
| Diarrhoea | 17/80 (21) | 25/80 (31) | 42/160 (26) |
| **Laboratory parameters** |  |  |  |
| Hyponatraemia (<130 mmol/L) | 17/78 (22) | 13/80 (16) | 30/158 (19) |
| Hypokalaemia (<3.0 mmol/L) | 9/78 (11.5) | 9/80 (11) | 18/158 (11) |
| Hypoglycaemia (< 3mmol/dl) | 4/80 (5) | 1/80 (1) | 5/160 (3) |
| Severe anaemia (Hb < 5g.dl) | 2/78 (3) | 1/79 (1) | 3/157 (2) |
| Lactate > 2 mmols/L | 39/68 (57) | 36/71 (51) | 75/139 (54) |
| Malaria film positive | 15/80 (19) | 7/80 (9) | 22/160 (14) |
| HIV Antibody positive | 1/80 (1) | 6 /80 (7.5) | 7/160 (4) |
| **Pre-existing Conditions and preadmission**<br>**Treatments** |  |  |  |
| Pulmonary Tuberculosis | 1/80 (1) | 2/80 (2.5) | 3/160 (2) |
| Congenital Heart Disease | 0 | 0 | 0 |
| Cerebral Palsy/severe developmental delay | 6/80 (7.5) | 3/80 (4) | 9/160 (6) |
| Currently taking antibiotics | 25/80 (31) | 28/80 (35) | 33/160 (21) |
| Currently taking antimalarials | 7/80 (9) | 9/80 (11) | 16/160 (10) |
| Currently taking antiretrovirals | 1/80 (1) | 6/80 (7.5) | 7/160 (4) |

Data are number (%) or median [interquartile range] unless otherwise specified.

### Adherence to randomised feed and volume received

Detailed summaries of feed volumes and adherence up to day 14 hospitalisation are reported in **Table S1**. Overall, ten children randomised to legume feeds were switched to WHO feeds. From hospital admission to day 14, 5 children switched to WHO feeds: 2 within 48-hours of admission as they could not tolerate non-liquid feeds or they developed a severe decompensation (2 of which died with severe decompensation) and 3 switched between days 3-13. Furthermore, 5 additional children switched feeds after day 14. Owing to the higher feed refusal in legume feed arm, the feed volumes given and percentage receiving the full amount were higher in the WHO arm on Day 0 and Day 1 however by Day 2 the total feed volume was similar between the two arms.

### Energy and protein intake

The daily summaries of energy and protein intake are reported in **Table S2**. Daily energy intake (reported in Kcals) was slightly higher in the WHO arm on Day 0 and Day 1 but was similar following this. Overall, energy intake met the nutritional target in both groups at all time points. Protein content in the legume feeds were much higher between Days 0-3 but was equivalent beyond this timepoint. In both arms children met expected protein intake targets at all time points.

### Length of Hospital Admission

71/80 (88.8%) and 67/80 (83.4%) for the legume feed and WHO feed respectively recovered and were discharged home. Their median hospital length was 11 days (IQR 7) and 12 days (IQR 8) respectively. The length of admission for those surviving to discharge, deaths and absconders are summarised in box-and-whiskers plots and table (**Supplemental Figure S1**). The fatal cases on the legume and WHO feeds arms had a median hospital stay of 6.5 days (IQR 4.75) and 13 days (IQR 9.5) respectively.

### Outcomes

Primary and secondary endpoints are summarised in **Table 2**. By intention to treat there was no difference in change in MUAC by day 90 (primary endpoint) with a median change in centimeters of 1.1 (1.1 IQR) and 1.4 (1.40 IQR) for the legume feed arm and WHO feeds arm respectively (p=0.09). Day 90 mortality (co-primary endpoint) for legume feed and WHO feeds arms were similar: 11/80 (13.8%) and 12/80 (15%) respectively (also see **Figure 2A** (Kaplan Meier for mortality by intention to treat). Most deaths occurred within the initial period of hospitalisation 7/11 (64%) and 8/12 (67%) respectively, from complications of infection predominantly associated with diarrhoea and pneumonia comorbidities (**Supplemental Table S3)**. As there were no differences in the baseline characteristics between arms in those treated per protocol (**Supplemental Table S4**) we conducted a per protocol analysis (which censored children who absconded during initial admission, those lost to follow-up Day 90 mortality and children who switched from legume feed to WHO feeds) for the primary and secondary endpoints. Per protocol there was little difference in the change in Day 90 MUAC from the intention to treat analysis. Day 90 mortality was lower in the legume feed arm 6/60 (10%) compared to 12/71 (17%) in the WHO arm but this was not statistically significant, hazard ratio 0.54 (95% confidence interval 0.20 – 1.45) p=0.22 (also see **Figure 2B**).

**Table 2.** Primary and Secondary Outcomes including safety outcomes.

|  | Legume | WHO | Statistic | p*-value |
| --- | --- | --- | --- | --- |
| **By <sup>a</sup>Intention to Treat** | N=80 | N=80 | Estimates (95% CI) |  |
| **Co-primary outcomes** |  |  |  |  |
| Median change (IQR) in MUAC Day 0 to Day 90 | 1.1 (1.1)cm | 1.4 (1.40)cm |  | 0.09* |
| Day 90 Mortality | 11/80 (14%) | 12/80 (15%) | 0.91 (0.40-2.07) | 0.83** |
| **Secondary Outcomes** |  |  |  |  |
| Day 90 Weight gains of >5g/kg/day | 5/69 (7%) | 5/68 (5%) |  | 0.45* |
| Day 90 Oedema resolution | 42/49 (86%) | 33/41 (81%) | 1.27 (0.80-2.00) | 0.29** |
| Day 90 Diarrhoea resolution | 12/17 (71%) | 20/25 (80%) | 0.57 (0.28–1.16) | 0.12** |
| Denovo development of diarrhoea (Day 2-14) | 16/80 (20%) | 18/80 (22.5%) | 0.84 (0.43-1.66) | 0.62** |
| Readmission to Day 90 | 2/80 (2.5%) | 4/80 (6%) | 0.50 (0.09-2.71) | 0.42*** |
| **<sup>b</sup>Per-protocol** | N=60 | N=71 |  |  |
| Median change (IQR) in MUAC Day 0 to Day 90 | 1.1 (1.30)cm | 1.4 (1.35)cm |  | 0.08* |
| Day 90 Mortality | 6/60 (10%) | 12/71 (17.5%) | 0.54 (0.20-1.45) | 0.22** |
| Day 90 Weight gain of >5g/kg/day | 5/64 (8%) | 5/68 (5%) |  | 0.53* |
| Day 90 Oedema resolution | 39/49 (79.5%) | 33/41 (81%) | 1.22 (0.82-2.07) | 0.27** |
| Day 90 Diarrhoea resolution | 12/15 (80%) | 20/25 (80%) | 0.57 (0.28- 1.17) | 0.12** |
| Denovo development of diarrhoea (Day 2-14) | 15/80 (18.8%) | 18/80 (22.5%) | 0.83 (0.42-1.64) | 0.59** |
| Readmission to Day 90 | 2/60 (3%) | 4/71 (6%) | 0.58 (0.11-3.14) | 0.53*** |

<sup>a</sup> **ITT Analysis:** Primary outcome results assessed based on their assigned randomised treatment (N=80), ignoring non-compliance with respect to the therapeutic feed intake.
<sup>b</sup> **PP Analysis:** Primary outcome results were assessed based on only the children who completed their originally allocated treatment.
\*p-value estimated from a *Mann-Whitney U* test
\*\*p-value estimated from unadjusted *Cox Regression* analysis
\*\*\* p-value represents Gray's test from a *Competing risk analysis, with mortality as the competing risk*

**Figure 2.**
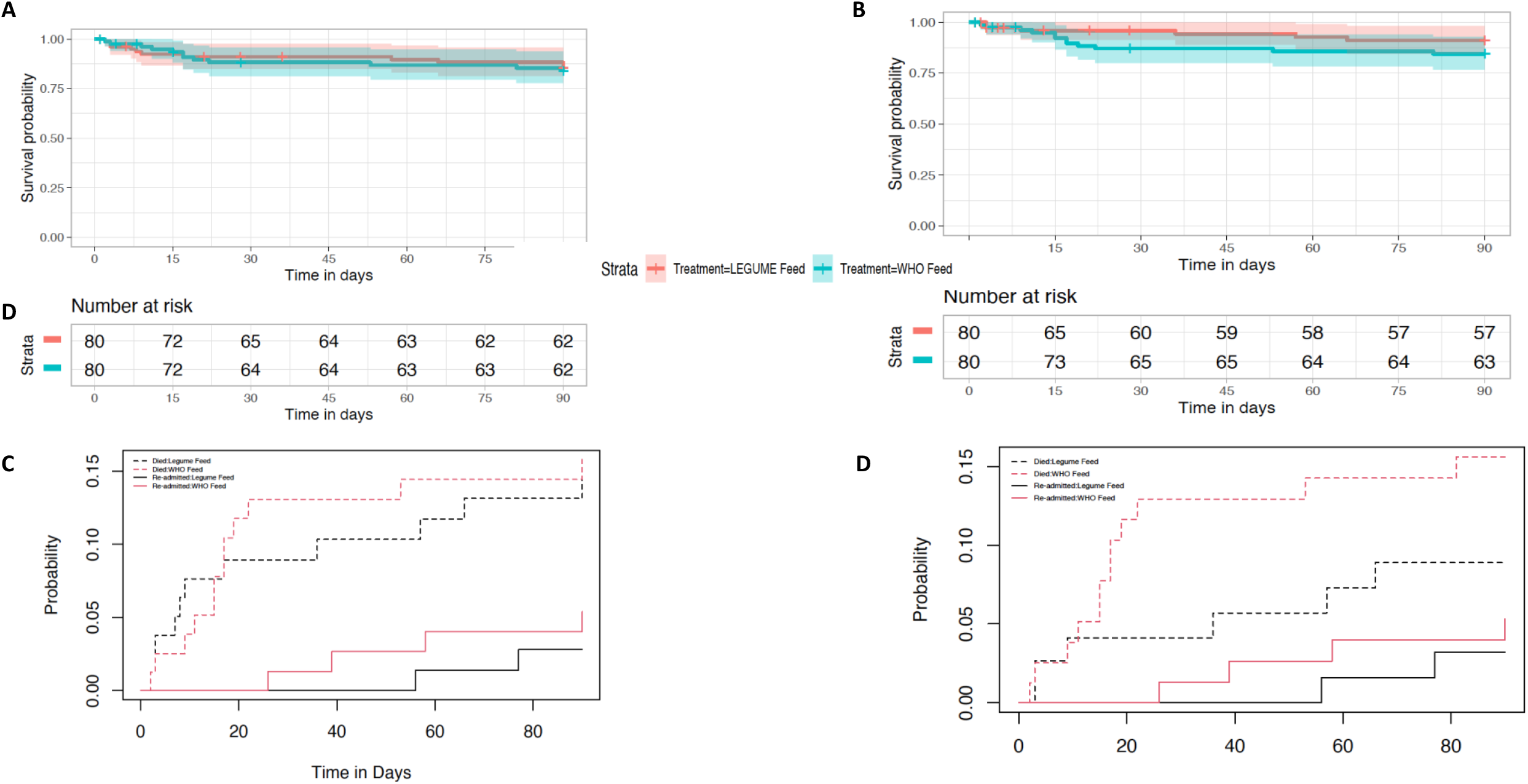
Survival and Readmission Plots to Day 90 A. **Figure 2A:** Kapan Meier plot with 95% confidence intervals from an ITT analysis. **B:** Kapan Meier plot with 95% confidence intervals from a PP analysis. **C**: Competing risk analysis curves of re-admissions with mortality as a competing risk from ITT analysis. **D:** Competing risk analysis curves of re-admissions with mortality as a competing risk from a PP analysis..

#### Secondary Endpoints

Few children achieved standard optimum of weight gain (>5g/kg) by day 90 in both arms, however the resolution of oedema (**Table 2**) was similar in both arms. Diarrhoea resolved in most children before Day 7 with no difference between arms. Development of denovo diarrhoea was high (34/160 (21.3%) overall with no difference between arms. With respect to serious adverse events, slightly more children in the WHO feeds arm were readmitted: 4 children (including one child twice) versus 2 in the legume feed arm (**Table 2**). The principal co-morbidities in the fatal cases mortality are summarised in **Table S3**. Readmission rates were assessed with competing risk analysis with mortality as a competing event. The analysis demonstrated that re-admissions were lower in the legume feed arm (3%) versus WHO feed arm (5%) in the ITT analysis Hazards ratio 0.50 (95% CI 0.09 – 2.71), p=0.42 and legume feed arm 2/69 (3%) versus WHO feed arm 4/71 (6%) in the PP analysis Hazard ratio 0.58 (95% CI 0.11 – 3.14) p=0.53 (**Figures 2 C and 2D**).

### Anthropometric changes in oedematous and non-oedematous children resolution

We were able to report detailed data on weight gain for study arms and for individual children overtime. Overall, mean (SD) MUAC and WHZ overtime are reported in **Figure 3**. In both study arms children transitioned from anthropometric parameters indicating severe malnutrition at trial entry to moderately and undernourished by Day 90. These parameters are summarised separately for oedematous and non-oedematous phenotypes (**Supplemental Figure S2)**. In addition, weight gain trajectory (gain, loss or maintenance) are reported over the follow up time period and stratified by presence of oedema at admission (**Table 3**) and for individuals in **Supplementary Figure S2**. We found little differences in early weight gains (to Day 7) in children without nutritional oedema. However, during this same period more children experienced weight loss in the legume arm in children presenting with oedema possibly due to the greater severity of oedema in the LF arm, which persisted to Day 28. By Day 90 weight gain occurred in 58/60 (97%) of children without oedema at baseline, whereas in children presenting with nutritional oedema 37/42 (88%) of the legume arm and 34/35 (97%) of the WHO arm had gained weight (p=0.613).

**Table 3.** The number of children with weight -gain, -loss and -maintenance based stratified by oedematous status at baseline **(as per ITT)**

| Time period | Non-Oedematous Children |  | Oedematous Children |  |
| --- | --- | --- | --- | --- |
| **Day 0- Day 7** | **WHO Feed** | **Legume Feed** | **WHO Feed** | **Legume Feed** |
| Weight Gain | 29/38 (76%) | 23/30 (77%) | 24/40 (60%) | 16/46 (35%) |
| Weight Loss | 5/38 (13%) | 4/30 (13%) | 13/40 (32.5%) | 25/46 (54%) |
| Maintenance | 4/38 (11%) | 3/30 (10%) | 3/40 (7.5%) | 5/46 (11%) |
| Statistic | 0.137 |  | 0.022 |  |
| **Day 7- Day 28** |  |  |  |  |
| Weight Gain | 21/35 (60%) | 20/29 (69%) | 27/36 (75%) | 25/44 (57%) |
| Weight Loss | 11/35 (31%) | 5/29 (17%) | 7/36 (19%) | 16/44 (36%) |
| Maintenance | 3/35 (9%) | 4/29(14%) | 2/36 (6%) | 3/44 (7%) |
| Statistic | 0.075 |  | 0.027 |  |
| **Day 28- Day 90** |  |  |  |  |
| Weight Gain | 24/33 (73%) | 23/27 (85%) | 28/35 (80%) | 34/42 (81%) |
| Weight Loss | 7/33 (21%) | 3/27 (11%) | 5/35 (14%) | 6/42 (14%) |
| Maintenance | 2/33 (6%) | 1/27 (4%) | 2/35 (6%) | 2/42 (5%) |
| Statistic | 0.865 |  | 0.101 |  |
| **Day 0- Day 90** |  |  |  |  |
| Weight Gain | 32/33(97%) | 26/27 (96%) | 34/35 (97%) | 37/42 (88%) |
| Weight Loss | 0/33 (0%) | 1/27 (4%) | 1/35 (3%) | 5/42 (12%) |
| Maintenance | 1/33 (3%) | 0/27 (0%) | 0/35 (0%) | 0/42 (0%) |
| Statistic | 0.824 |  | 0.613 |  |

\*p-value: represents Mann-Whitney U statistical analysis
**Weight gain:** Weight<sub>tB-tA</sub>>0
**Weight maintenance:** Weight<sub>tB-tA</sub>= 0
**Weight loss:** Weight<sub>tB-tA</sub> <0

**Figure 3.**
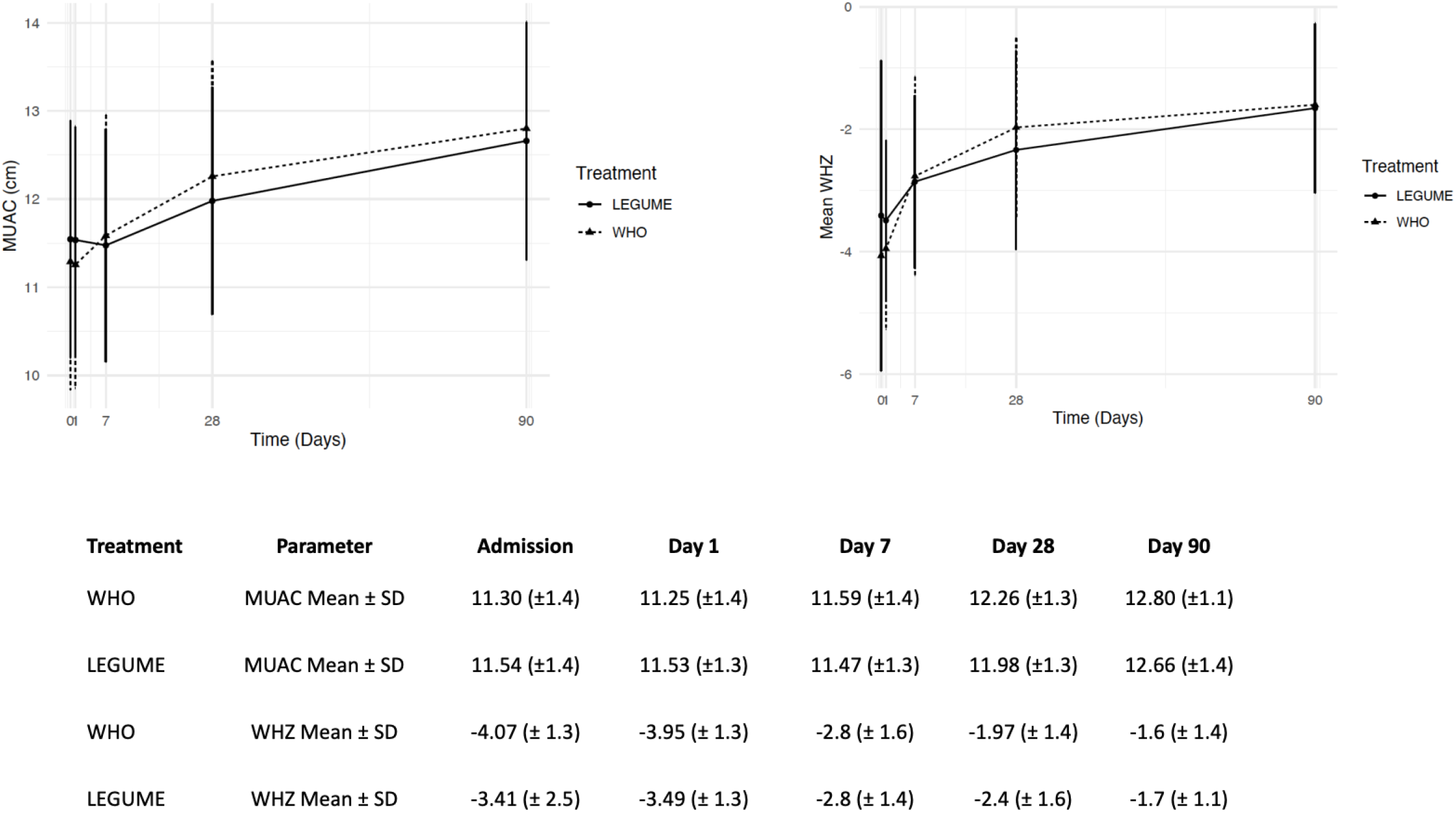
Mean (Standard Deviation) of mid-upper arm circumference (MUAC) and weight for height Z score (WHZ) from admission to Day 90.

## Discussion

In this trial comparing two nutritional strategies in 160 Ugandan children admitted with severe acute malnutrition, including 57% with the kwashiorkor phenotype, we were able to demonstrate that legume-enriched feed provided similar anthropometric outcomes than children receiving the milk-based WHO formulae (F75 followed by F100). In general, the weight gain velocities were less than the recommended > 5g/kg/day for both strategies with oedematous children experiencing much lower growth velocities. By Day 90 most childrens’ anthropometric parameters were consistant with moderate to mild undernutrition. Mortality remained high, overall 14% (23 children) died within 20 days of admission which were largely to due the complications of underlying infections (pneumonia and diarrhoea). In the intention to treat analyses we found no evidence for a difference in mortality between arms. In a per protocol analysis, we found Day 90 mortality was lower in the legume feed arm (10%) versus the standard feed arm (17.5%) and the rate of readmission by 90 days was less, 3% versus 5% respectively but neither of these finding were significant owing to the small sample size.

This trial was not directly powered to find a difference between the legume feed strategy and WHO feeds on patient-centred outcomes including de novo development of diarrhoea, mortality or readmissions. However, we did show that this novel strategy provided similar anthropometric improvements to the WHO feed arm and with no evidence of harm. Owing to poor palatability of the nutritional paste (children preferred liquid-based feeds initially) a number of children switched early to WHO feeds which has implications for the future design of other legume based feeds. On measured factors at baseline the characteristics remain balanced between the two arms in the per protocol analysis population, indicating none or minimal sampling bias from restricting to this population (**Supplemental Table S4**).

Whilst we considered resolution of diarrhoea as an endpoint, the accuracy of the reporting of this is the least robust outcome measure for a clinical trial. Parental reports of diarrhoea (defined as more than three loose stools) masked the spectrum of severity even though this largely resolved within a few days. However, as we have reported previously,^27^ another 23% of children developed de novo diarrhoea by two weeks whilst receiving nutritional rehabilitation. Children with SM hospitalised with diarrhoea and those developing diarrhoea are at risk of worse outcomes^27^ as well children with uncomplicated SM with diarrhoea managed in the community^27 28^. In this trial we found that the most common clinical complication contributing to in-patient deaths was diarrhoea in the WHO arm (5 patients) versus 1 patient in the legume feed arm. Current WHO guidelines indicates that diarrhoea is a trivial consequence of severe malnutrition^29^, however emerging data indicates that there is substantial evidence of profound gut-barrier dysfunction, characterised by blunted villi^30^, inflammation and increased permeability^31^. In addition, children with SM often having functional lactase, maltase and sucrase deficiency (the key F75/F100 disaccharides), which combined will exacerbate diarrhoea, impair vital nutritient uptake and impair recovery. As a result current formula have been adapted to reduce the sucrose load by incorporation of malodextran, which has a low risk of causing osmotic diarrhoea. Attempts to modify the initial starter feed (F75) by reducing lactose and carbohydrate-load. failed to improve outcomes (including time to stabilisation, diarrhoea and mortality). A further trial initially provided elemental feeds (hypoallergenic and anti-inflammatory feeds) but this did not improve markers biomarkers of intestinal and systemic inflammation and mucosal integrity^32^. This indicates more radical revisions to the formula are required^5^. We had proposed that a lactose-free, fermentable carbohydrate-containing (chickpea) alternative^22^ may address the poor outcomes in this high risk group.

Progress in the area of optimal nutritional feed for those who have been hospitalised with SM has been very slow and piecemeal. Most field research conducted in Africa is largely in community-based programmes (uncomplicated SM) often with good outcomes. Future research investigating whether innovative feeding strategies focusing on gut repair, optimizing the microbial environment as well as providing nutritional support after immediate recovery, could improve clinical outcomes compared to standard treatments (and less costly). This would be a substantive starting point to revise treatment guidelines. With respect to availability most nutritional feeds are largely manufactured remote from the continent or the communities mostly affected. Feed availability is dependent upon the international donors, at substantial costs, thus accessibility for local communities is low^33^. International non-governmental organisations have recognised that there is an unmet need and to develop them more locally as current formulations for inpatient and community feeds require dried milk, which is often limited, variable in quality and expensive.

This trial was the first step in providing some evidence that food products, which are all available locally in Uganda, could be used in future feed designs to address this unmet need directly and the research gap highlighted in the WHO report on RUTF feed composition^34^ Similar consultations for reviewing the content of inpatient feeds are lacking.

## Data Availability

All data produced in the present study are available upon reasonable request to the authors

## Contributors

Conceptualisation : GF, KM and KW; Formal Analysis: AK and ECG; Investigation: POO, WO, TS, CBO, RM, AM and KM Data Curation: AK and KW; Writing – Review & Editing: AK, KW, KM, GF,ECG and POO

All authors vouch for the completeness and accuracy of the data and analyses presented.

## Acknowledgements

We thank all the participants and staff from all the centres participating in the MIMBLE trial. We would like to thank Siraj Kijogo (Head) and Jennifer Adong (Senior Nurse) of the Mbale Regional Referral Hospital Nutrition Unit.

G.F. is an NIHR senior investigator. This study is funded by an award to KM and GF UK by **Imperial** Confidence in Concept – Joint Translational Fund. Support for the trial management [KEMRI Wellcome Trust Programme East African Overseas Programme Award (2016) from the Wellcome Trust 203077/Z/16/Z])

## Data sharing Statement

The datasets generated during the trial will be available upon reasonable request, following the publication of the trial results, from Prof. Gary Frost. Anonymised data including clinical and anthropometric data will be made available. The data used in this research was collected subject to the informed consent of the participants. Access to the data will only be granted in line with that consent, subject to approval by the project ethics board and under a formal Data Sharing Agreement.

