## Supplemental File for "Modifying gut integrity and microbiome in children with severe acute malnutrition using legume-based feeds (MIMBLE II): A Phase II trial"

**Supplemental Tables and Figures**

**Table S1** Daily feed Provision for children receiving standard feed (WHO) and legume based feeds (LF)
**Table S2** Daily energy (in kilocalories) and protein (in grams) and proportion meeting target intakes
**Table S3** Mortality in children summarising underlying complications/conditions
**Table S4** Baseline characteristics comparisons for WHO and legume feeds for intention to treat (ITT) and per protocol (PP) analysis.
**Table S5** Multiple imputations (MI) analysis Table
**Figure S1** Status of children and length of hospital stay by study arm

**Figure S2** Individual weight line plots based on oedematous status from baseline

**Table S1** Daily feed Provision for children receiving standard feed (WHO) and legume-based feeds (LF)

| | n/N (%) that Received any feed | | Total N of Deceased Participants | | γTotal N switched to WHO feed | n/N (%) Received fully prescribed feed | | Chi-square p-value |
|---|---|---|---|---|---|---|---|---|
| Days | WHO | LF | WHO | LF | Legume Group only | WHO | LF | |
| 0 | 80/80 (100) | *78/80 (97.5) | 0 | 0 | 0 | 61/80 (76.25) | 36/78 (46.1) | <0.001 |
| 1 | **79/80 (98.75) | ^78/80 (97.5) | 0 | 0 | 1 | 72/79 (91.1) | 56/78 (71.8) | 0.003 |
| 2 | 79/80 (98.75) | α72/79 (91.1) | 0 | 1 | 4 | 69/79 (87.3) | 53/72 (73.6) | 0.053 |
| 3 | 78/79 (98.7) | 70/77 (90.9) | 1 | 3 | 5 | 65/78 (83.3 | 53/70 (75.7) | 0.344 |
| 4 | ***75/78 (96.1) | 69/77 (89.6) | 2 | 3 | 6 | 66/75 (88.0) | 59/69 (85.5) | 0.845 |
| 5 | ~73/78 (93.6) | 68/77 (88.3) | 2 | 3 | 7 | 65/73 (89) | 56/68 (82.35) | 0.370 |
| 6 | 73/78 (93.6) | 68/76 (89.5) | 2 | 4 | 7 | 64/73 (87.7) | 51/68 (75) | 0.085 |
| 7 | 70/74 (94.6) | β66/75 (88) | 2 | 5 | 6 | 60/70 (85.7) | 49/66 (74.2) | 0.144 |
| 8 | 62/66 (93.9) | 55/65 (84.6) | 2 | 5 | 7 | 47/62 (75.8) | 43/55 (78.2) | 0.933 |
| 9 | #50/55 (90.9) | 52/62 (83.8) | 2 | 5 | 6 | 41/50 (77.4) | 40/52 (76.9) | 0.697 |
| 10 | 43/48 (89.5) | 45/55 (81.8) | 3 | 5 | 6 | 34/43 (79.1) | 35/46 (76.1) | 1 |
| 11 | 35/40 (87.5) | 40/49 (81.6) | 3 | 5 | 5 | 27/35 (90) | 25/40 (62.5) | 0.262 |
| 12 | 30/35 (85.7) | 32/42 (76.1) | 4 | 5 | 4 | 25/30 (83.3) | 25/32 (78.1) | 0.844 |
| 13 | 27/32 (84.3) | 31/42 (73.8) | 4 | 5 | 5 | 22/27 (81.5) | 23/31 (74.2) | 0.728 |
| 14 | 24/29 (82.7) | 26/39 (66.6) | 4 | 5 | 5 | 21/24 (87.5) | 19/26 (73.1) | 0.358 |
| Notes | *2 children refused feed on admission and one out of the two later on was started on F75 on day 1 and died the next day<br>** Child absconded from hospital<br>***2 Children absconded from hospital.<br>~ 1 child absconded.<br># 1 child absconded.<br>^ 1 child absconded.<br>α 1 child absconded.<br>β 1 child absconded.<br>γ The number varies with deaths occurring on children after they switched feeds. | | | | | | | |

**Table S2.** Daily energy (in kilocalories) and protein (in grams) and proportion meeting target intakes

| | **Median (IQR) of Energy Intake in Kcal** | | **Median (IQR) Intake of energy vs target %** | | **Mann Whitney U (p-value)** | **Median (IQR) of Protein Intake in g** | | **Median of protein intake compared to target %** | | **Mann Whitney U (p-value)** |
|---|---|---|---|---|---|---|---|---|---|---|
| Days | WHO | LF | WHO | LF | Between Treatments | WHO | LF | WHO | LF | Between Treatments |
| 0 | 553.1 (208.13) | 499.8 (235.2) | 100 (0) | 95.6 (24.4) | <0.001 | 6.6 (2.5) | 13.8 (6.5) | 100 (0) | 95.6 (24.4) | <0.001 |
| 1 | 563 (225) | 592.3 (214.8) | 100 (0) | 100 (2) | 0.002 | 6.8 (2.7) | 16.5 (6) | 100 (0) | 100 (2) | 0.002 |
| 2 | 585 (247) | 609.6 (203.6) | 100 (0) | 100 (1.2) | 0.007 | 6.8 (2.7) | 16.8 (8.3) | 100 (0) | 100 (1.2) | 0.007 |
| 3 | 660 (267.75) | 643.1 (363.7) | 100 (0) | 100 (0.5) | 0.171 | 13.1 (16.2) | 17.7 (10) | 100 (0) | 100 (0.5) | 0.171 |
| 4 | 730 (287) | 670.6 (415.5) | 100 (0) | 100 (0) | 0.411 | 20.9 (18.2) | 18.5 (11.4) | 100 (0) | 100 (0) | 0.411 |
| 5 | 780 (376.7) | 713.2 (303.3) | 100 (0) | 100 (0) | 0.138 | 22.6 (19.5) | 19.6 (8.4) | 100 (0) | 100 (0) | 0.138 |
| 6 | 810 (337.5) | 731.5 (397.7) | 100 (0) | 100 (0.9) | 0.063 | 23.5 (10.5) | 20.2 (10.9) | 100 (0) | 100 (0.9) | 0.064 |
| 7 | 840 (333.7) | 783.3 (458.2) | 100 (0) | 100 (1.1) | 0.228 | 24.4 (12.7) | 21.6 (12.6) | 100 (0) | 100 (1.1) | 0.228 |
| 8 | 810 (380.6) | 853 (477.5) | 100 (0.4) | 100 (0) | 0.385 | 23.1 (19.1) | 23.5 (13.2) | 100 (0.4) | 100 (0) | 0.385 |
| 9 | 836.5 (397) | 853.4 (548.6) | 100 (0) | 100 (0) | 0.538 | 23.9 (20.3) | 22.5 (14.9) | 100 (0) | 100 (0) | 0.538 |
| 10 | 810 (397) | 975.4 (601) | 100 (0) | 100 (0) | 0.876 | 23.5 (20.2) | 26.9 (16.6) | 100 (0) | 100 (0) | 0.876 |
| 11 | 840 (339) | 817.8 (548.1) | 100 (0) | 100 (8) | 0.196 | 24.4 (19.4) | 22.5 (15.1) | 100 (0) | 100 (8) | 0.196 |
| 12 | 885 (391.7) | 877.8 (603.5) | 100 (0) | 100 (0) | 0.560 | 25.2 (17.2) | 24.2 (16.6) | 100 (0) | 100 (0) | 0.560 |
| 13 | 870 (397) | 853.4 (694.9) | 100 (0) | 100 (1) | 0.468 | 23.5 (21) | 23.5 (19.1) | 100 (0) | 100 (1) | 0.468 |
| 14 | 851 (399.2) | 829 (521.2) | 100 (0) | 100 (4) | 0.007 | 19.4 (20.2) | 22.8 (14.4) | 100 (0) | 100 (4) | 0.007 |
| Notes | *2 children refused feed on admission and one out of the two later on was started on F75 on day 5 up to day 12 of discharge.<br>** Child absconded from hospital.<br>***2 Children absconded from hospital.<br>~ 1 child absconded.<br># 1 child absconded.<br>^ 1 child absconded.<br>α 1 child absconded.<br>β 1 child absconded. | | | | | | | | | |

**Table S3** Mortality in children summarising underlying complications/conditions

| **Treatment** | WHO | LF | Comment |
|---|---|---|---|
| **Mortality All** | 12 | 11 | |
| *Inpatient deaths* | 8 | 7 | |
| *Major clinical syndrome associated with inpatient death* | | | |
| Diarrhoea | 5 | 1 | |
| LRTI | 1 | 4 | |
| Sepsis other | 1 (Measles) | 1 (malaria) | |
| Tuberculosis | 1 | | |
| Other | | 1 | Pancytopenia |
| Post discharge deaths | 4 | 4 | |
| (HIV-related) | (3) | (1) | |
| **Readmissions** | | | |
| Readmission and death | 2 | 1 | |
| Readmission only | 2 | 1 | |

**Post discharge deaths WHO arm**:
Readmission/death: 1 diarrhoea 1 sepsis; 2 deaths in community unknown
**Post discharge deaths Legume feed**:
Readmission/death 1 diarrhoea; Deaths in community: 1 diarrhoea, relapse of kwashiorkor, measles

**Table S4 Baseline characteristics comparisons for WHO and legume feeds for intention to treat (ITT) and per protocol (PP) analysis.**

| **Characteristic** | By intenstionto treat **Per-**protocol | | **Per-protocol Analysis** | |
|---|---|---|---|---|
| | **Legume feed** | **WHO feeds (F75/F100)** | **Legume feed** | **WHO feeds (F75/F100)** |
| Participants, n | 80 | 80 | 60 | 71 |
| Median age in months [Interquartile range) | 18 [12.7) | 17 [11.7] | 17 [12.2] | 17 [11.5] |
| Sex: Male (%) | 44 (55) | 39 (48.75) | 33 (51.6) | 34 (53.12) |
| **Nutritional status and history** | | | | |
| Median mid-upper arm circumference, cm [IQR] | 11.4 [1.7] | 11.2 [1.7] | 11.4 [1.32] | 11.2 [1.8] |
| Weight-for-height/length z score <-3 | 31 (39) | 39 (49) | 23 (38) | 39 (49) |
| Weight for height Z score [IQR] | -3.79 [1.7] | -4.08 [3] | -4.05 [1.57] | -4.19 [2.08] |
| Oedema (kwashiorkor) | 49 (61) | 41 (51) | 37 (62) | 39 (44) |
| Severe/generalized Oedema | 10/49 (20) | 6/41 (15) | 6/37 (16) | 6/39 (15) |
| Desquamation or flaky paint skin | 20/80 (25) | 15/80 (19) | 14/60 (23) | 16/71 (22.5) |
| Age when feeds introduced (months) | 4 [3] | 5 [3] | 4 [3] | 5 [2.5] |
| Currently breast feeding | 21/80 (26.5) | 24/80 (30) | 15/60 (25) | 21/71 (30) |
| Previous admission with SAM | 5/80 (6) | 4/80 (5) | 4/60 (7) | 4/71 (6) |
| **Complications at Presentation** | | | | |
| History of fever | 63/80 (79) | 57/80 (71) | 48/60 (80) | 50/71 (70) |
| Fever (axillary temp) > 37.5ºC | 10/80 (12.5) | 10/80 (12.5) | 9/60 (15) | 8/71 (11) |
| Cough | 59/80 (74) | 59/80 (74) | 45/60 (75) | 54/71 (76) |
| Indrawing or deep breathing | 3/80 (4) | 3/80 (4) | 2/60 (3) | 3/71 (4) |
| Vomiting | 23/80 (29) | 25/80 (31) | 13/60 (22) | 30/71 (30) |
| Diarrhoea | 17/80 (21) | 25/80 (31) | 11/60 (18) | 19/71 (27) |
| **Laboratory parameters** | | | | |
| Hyponatraemia (<130 mmol/L) | 17/78 (22) | 13/80 (16) | 16/60 (27) | 11/71 (15.5) |
| Hypokalaemia (<3.0 mmol/L) | 9/78 (11.5) | 9/80 (11) | 6/60 (10) | 8/71 (11) |
| Hypoglycaemia (< 3mmol/dl) | 5/80 (6) | 1/80 (1) | 3/60 (5) | 0 |
| Severe anaemia (Hb < 5g.dl) | 2/78 (3) | 1/79 (1) | 1/60 (2) | 1/71 (1) |
| Lactate > 2 mmols/L | 39/68 (57) | 36/71 (51) | 22/60 (37) | 28/71 (39) |
| Malaria film positive | 15/80 (19) | 7/80 (9) | 12/60 (20) | 6/71 (8.5) |
| HIV Antibody positive | 1/80 (1) | 6 /80 (7.5) | 1/60 (2) | 5 /71 (7) |
| **Pre-existing Conditions /preadmission treatments** | | | | |
| Pulmonary Tuberculosis | 1/80 (1) | 2/80 (2.5) | 1/60 (2) | 2/71 (3) |
| Congenital Heart Disease | 0 | 0 | 0 | 0 |
| Cerebral Palsy/severe developmental delay | 6/80 (7.5) | 3/80 (4) | 4/60 (7) | 3/71 (4) |
| Currently taking antibiotics | 25/80 (31) | 28/80 (35) | 19/60 (32) | 26/71 (37) |
| Currently taking antimalarials | 7/80 (9) | 9/80 (11) | 7/60 (12) | 7/71 (10) |
| Currently taking antiretrovirals | 1/80 (1) | 6/80 (7.5) | 1/60 (2) | 5/71 (7) |

Data are number (%) or median [interquartile range] unless otherwise specified.

$^{\alpha}$ **ITT Analysis:** Primary outcome results assessed based on their assigned randomised treatment (N=80), ignoring non-compliance with respect to the therapeutic feed intake. $^{\beta}$ **PP Analysis:** Primary outcome results were assessed based on only the children.

**Table S5** Multiple Imputation listing

Number of Multiple Imputation Analysis values on MUAC, Weight & Height, Oedema and Diarrhoea

| Total N of Imputed Missing Data by Treatment by Time-point | | | | | |
|---|---|---|---|---|---|
| Treatment | **D0** | **D1** | **D7** | **D28** | **D90** |
| **WHO** | 0 | 1 | 3 | 5 | 9 |
| **Legume** | 0 | 0 | 2 | 7 | 11 |

***Treatment, Weight, Height and MUAC diarrhoea and oedema at baseline were used in the model as complete variables.**

**Figure S1** Status of children  and length of hospital stay by study arm.

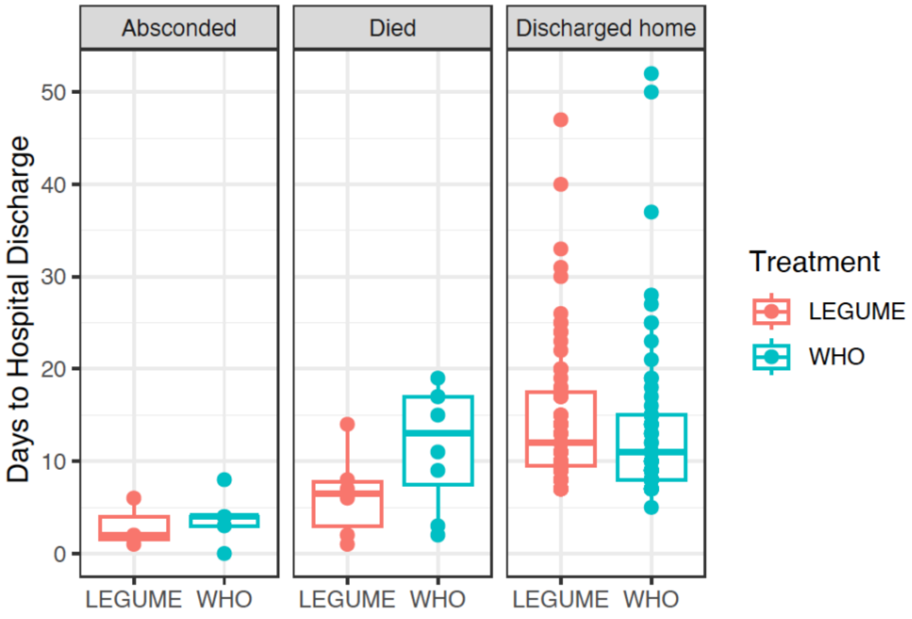


| Treatment | Status | N | Median | IQR |
|---|---|---|---|---|
| LEGUME | Absconded | 3 | 2 | 2.5 |
| LEGUME | Died | 6 | 6.5 | 4.75 |
| LEGUME | Discharged home | 71 | 12 | 8 |
| WHO | Absconded | 5 | 4 | 1 |
| WHO | Died | 8 | 13 | 9.5 |
| WHO | Discharged home | 67 | 11 | 7 |

**Figure S2** Individual weight line plots based on oedematous status from baseline.

**a)** Represents individual weight between treatments across time (days) of children that presented with no oedema upon admission.

**b)** Represents individual weight between treatments across time (days) of children that presented with oedema upon admission.

Black lines represents the mean.

**a**) Non-oedematous at baseline

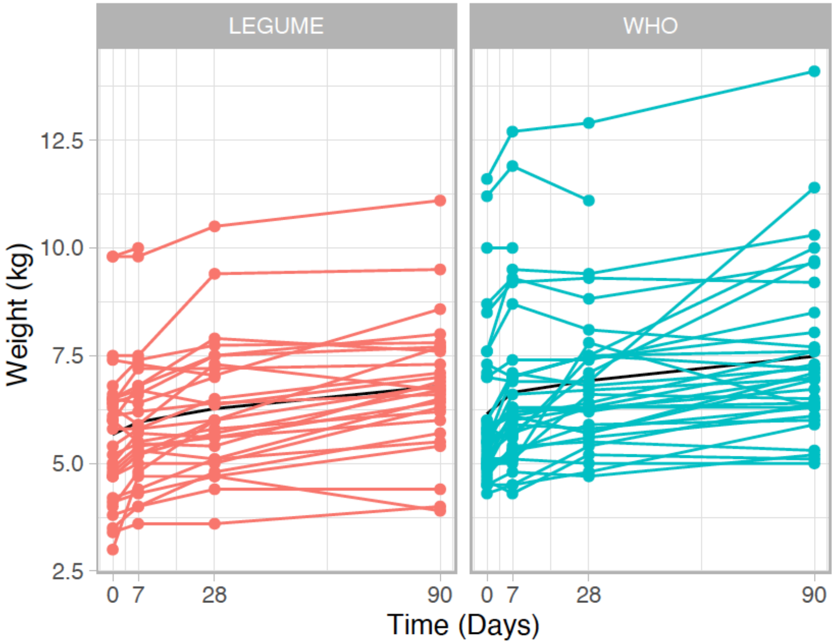


**b)** Oedema present at baseline

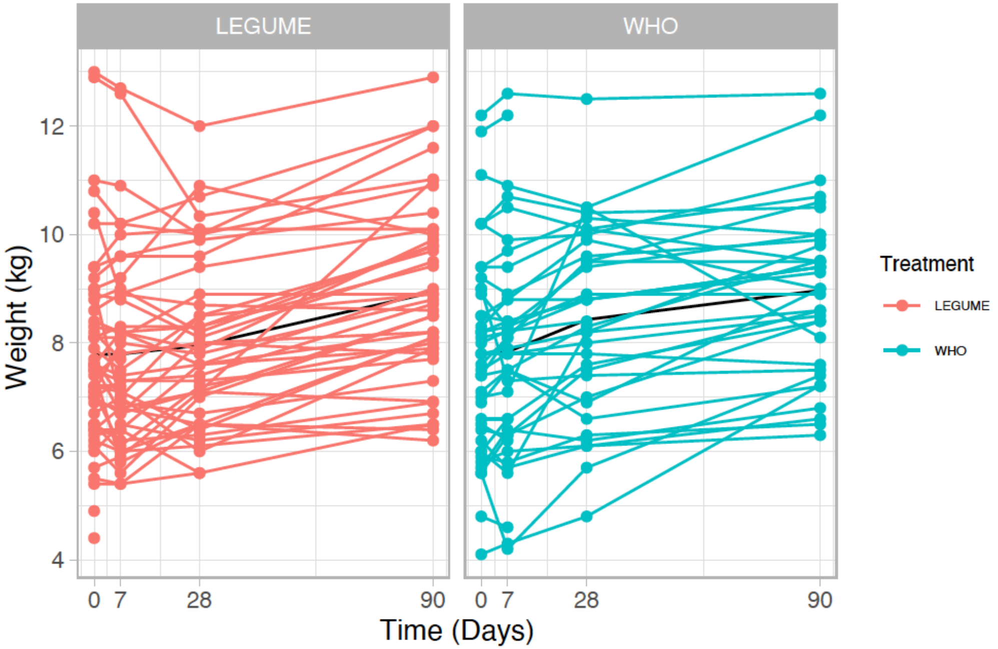
